# Sustaining Social Distancing Policies to Prevent a Dangerous Second Peak of COVID-19 Outbreak

**DOI:** 10.1101/2020.04.17.20069351

**Authors:** Zhilan Feng, Haiyun Damon-Feng, Henry Zhao

## Abstract

Governments around the world have enacted strict social distancing policies in order to slow the spread of COVID-19. The next step is figuring out when to relax these restrictions and to what degree. Our results predict potentially disastrous implications of ending these policies too soon, based on projections made from a Susceptible-Exposed-Infectious-Removed (SEIR) epidemic model. Even when infection rates appear to be slowing down or decreasing, prematurely returning to “business as usual” produces a severe second peak far worse than the first. Furthermore, such a second peak is made more likely when very severe restrictions are initially enacted. Only an appropriately measured and committed set of restrictions can appropriately control COVID-19 outbreak levels.

## Introduction

As countries around the world navigate the first month of social distancing restrictions, the questions of when these policies can be relaxed and to what degree will become fundamentally important in constructing policy responses to COVID-19 control moving forward. Since these policies are socially and economically taxing, the public as well as policymakers will generally want them to end, particularly when infection rates appear to have plateaued or even begun to decrease. These rollbacks in restrictions are already being discussed in countries around the world. However, relaxing these restrictions too soon can have dire consequences if it ultimately allows COVID-19 to spread even more dangerously than it did before. We use a standard SEIR epidemic model incorporating the most up-to-date estimates for COVID-19 relevant parameters in order to explore the implications of a potentially catastrophic second peak of the outbreak in the wake of premature declarations of success. This report illustrates several informative examples that highlight the most important takeaway: social distancing policies must be sustained even in the face of declining infection rates, or the entire effort to contain COVID-19 fails. An interactive notebook created using *Mathematica* featuring our results can be accessed at the following link: https://bit.ly/2yY9dnM.

## Methods

We investigate the possible consequences of relaxing the strict social distancing policies by examining the solution behaviors of a Susceptible-Exposed-Infectious-Removed (SEIR) type of epidemic model. (See ref. (*1*) for more explanations about this type of models). Our model accounts for both symptomatic and asymptomatic people (a critically important category for COVID-19 disease spread). The population is divided into five epidemiological classes: susceptible (*S*), exposed (*E*), asymptomatic (*A*), and removed (*R*). The total population is *N* = *S* + *E* + *A* + *I* + *R*. The model consists of the following differential equations:

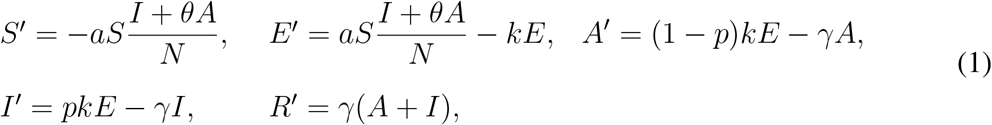

with initial conditions: *S*(0) = *N −I*_0_, *I*(0) = *I*_0_ *>* 0, *E*(0) = *A*(0) = *R*(0) = 0. It is assumed in the model that among the infectious people, proportions *p* and 1 *− p* are symptomatic and asymptomatic, respectively. Asymptomatic individuals can also transmit the disease but at a lower rate than symptomatic individuals, which is denoted by the factor *θ <* 1. The mean latent and infectious periods are 1*/k* and 1*/γ*, respectively. The parameter *a* denotes the *per capita* effective contact rate, i.e., contacts that can lead to infection. In the case of no intervention, *a* = *a*_0_ = *ℛ*_0_*γ*, where *ℛ*_0_ is the basic reproduction number. For ease of reference, we will refer to *a* simply as contact rate. The key effect of social distancing policies is to reduce the contact rate, We assume that *a*(*t*) is a function of time and its values are different before and after the strict intervention policies are lifted. We consider a step function defined as

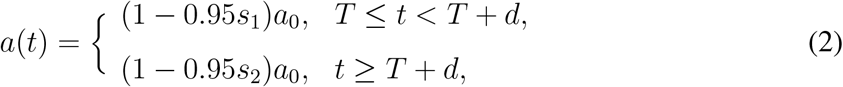

where *T* denotes the time when the lockdown order starts and *d* represents the duration of the order, while *s*_1_ denotes the reduction of contacts during initial and after loosed restrictions, respectively, both from business as usual (0) to no contact (1). We will explore the scenarios by simulating the model with various combinations of parameters *s*_1_ and *s*_2_. For the simulation results in Figures 1 and 2, we choose the parameter values to be suitable for COVID-19 as the follows: 1*/k* = 10 days, 1*/γ* = 7 days, *p* = 0.8, *θ* = 0.5, and *ℛ*_0_ = 5. It has recently been reported (see ref. (*2*)) that *ℛ*_0_ = 5.7, which is about twice as high as the commonly cited values of 2 to 3, making it even more challenging to prevent a second wave of this outbreak with a high peak.

**Figure 1.**
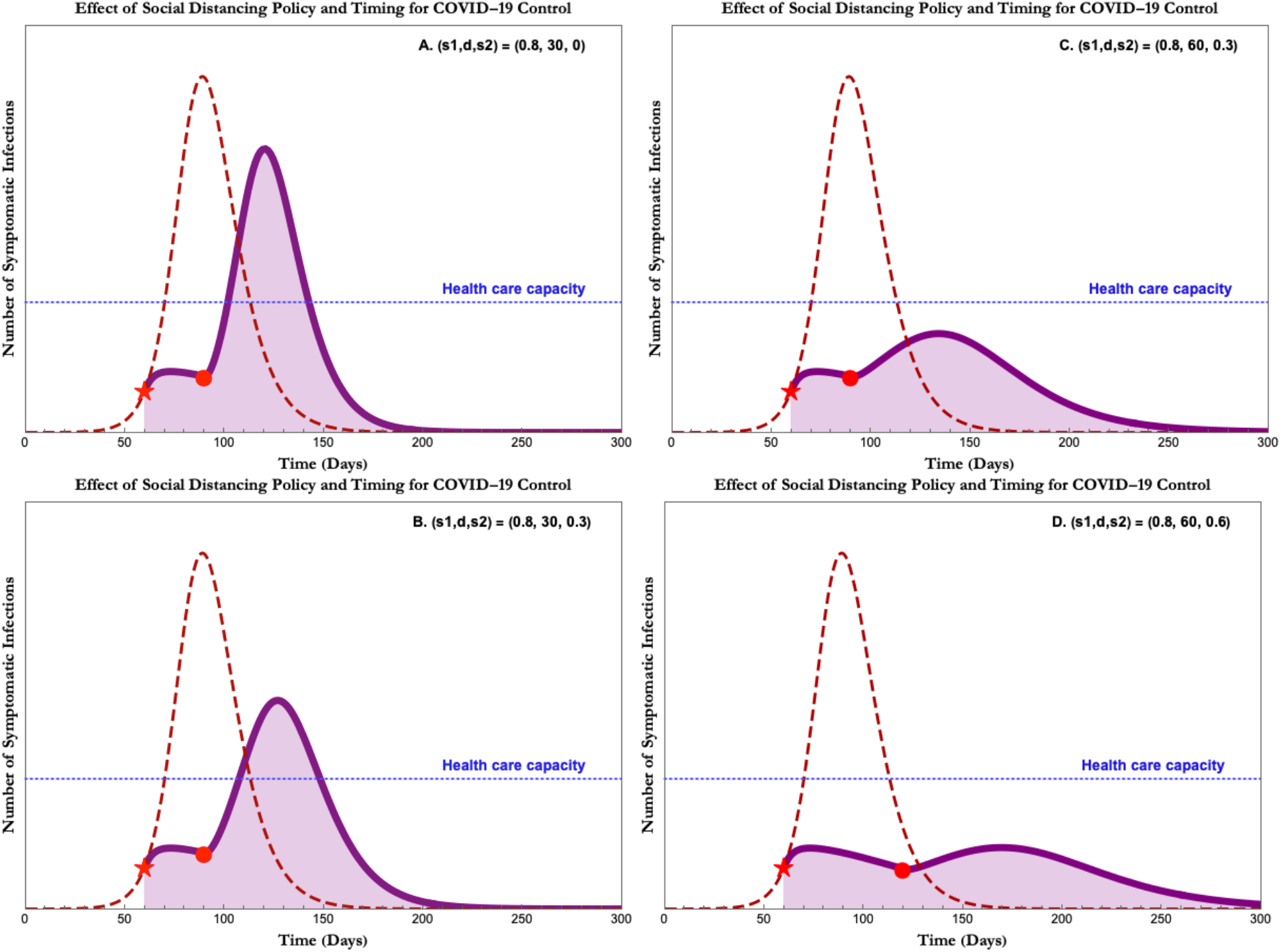
Projected outbreak of COVID-19 given intervention parameters. In case (A), restrictions are completely lifted after thirty days. In case (B), some residual restrictions are kept in place but restrictions are mostly lifted after thirty days. In case (C), these residual restrictions are kept in place after sixty days of the initial restrictions. In case (D), stronger residual restrictions are kept in place after sixty days. The dashed red curve represents the projected outbreak of COVID-19 without any intervention, while the solid purple curve represents the projected outbreak under a particular intervention strategy. These strategies consist of the severity (s_1_) and duration (d) of initial restrictions, as well as the severity of residual restrictions (s_2_) in place after the initial period.

**Figure 2.**
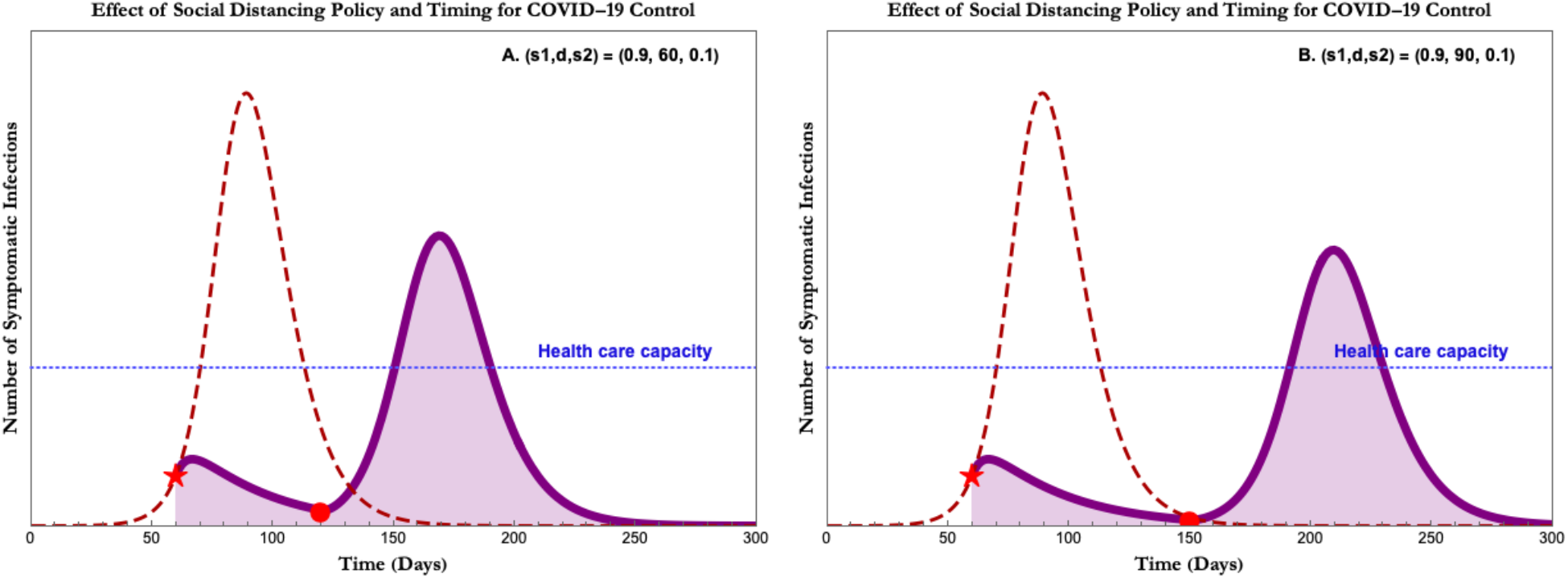
Projected outbreak of COVID-19 given intervention parameters. Similar to Figure 1 except initial severity s_1_=0.9. In case (A), restrictions are almost entirely lifted after sixty days. In case (B), the initial restrictions are kept in place for ninety days. In both cases, a dangerously high second peak exists despite infection rates trending toward zero.

## Results

Figures 1 and 2 summarize the main implications of our results. In each individual plot, the dashed red curve represents the projected outbreak of COVID-19 without any intervention, while the solid purple curve represents the projected outbreak under a particular intervention strategy. These strategies consist of the severity (*s*_1_) and duration (*d*) of initial restrictions, as well as the severity of residual restrictions (*s*_2_) in place after the initial period. For context, the stay-at-home order issued in the United States would correspond to an initial severity *s*_1_ = 0.8 and *d* = 30, while even stricter policies such as Italy’s lockdown would correspond to a higher severity of *s*_1_ = 0.9. A return to business as usual corresponds to *s*_2_ = 0, while common sense restrictions such as banning all large-scale public events corresponds to *s*_2_ = 0.3. The particular projections presented in these figures correspond to previous policy choices and options going forward, as discussed below.

### Consequences of loosening restrictions too soon: 30 days

While lockdown and shelter-in-place protocols would likely be effective at preventing infection rates from skyrocketing above health care capacity, we predict that returning to business as usual too soon after seeing preliminary results - including results indicating that the infection rates have slowed and that the outbreak has “peaked” - will have disastrous consequences (see Figure 1A). Returning to business as usual prematurely will likely result in a severe secondary outbreak. This is due in part to the longer-term total epidemiological cycle of COVID-19. If social distancing policies are lifted shortly after the peak, there are still a high number of infectious people and a high proportion of the population remains susceptible to infection. Lifting or relaxing social distancing policies at this point in the epidemiological cycle would allow COVID-19 to spread again and cause a severe second peak that would either require renewed intervention or cause a severe second peak, overwhelming the health care system and leading to more deaths. Even if some residual restrictions remain in place, such as cancellations of large social events, this secondary outbreak will likely still spike above health care capacity (see Figure 1B). Our model suggests that if restrictions are lifted or relaxed after 30 days, even if infection rates appear to be decreasing at that point, the restrictions should be only marginally relaxed. Otherwise, there will likely be a second outbreak peak.

### Consequences of loosening restrictions too soon: 60 days

Doubling the duration of severe social distancing restrictions to 60 days yields more stability with respect to managing a severe second peak. However, significant relaxation of restrictions after 60 days (even with some “common sense” restrictions like limiting extremely large scale gatherings) is likely to result in a second spike of COVID-19 infections that would again strain, and possibly exceed, health care capacity (see Figure 1C). However, if restrictions are only somewhat loosened after 60 days, the results would likely be much more manageable. There would continue to be infections and a resulting second peak, but a much “flatter” peak than under the 30-day scenario (see Figure 1D). An additional consideration that is not reflected in our model is that the health care capacity may not be static throughout the pandemic. We know that health care resources are being utilized and depleted, and it is possible that the health care system’s capacity in 60 days will be lower than it was at the beginning of the pandemic. This underscores the importance of maintaining social distancing policies to prevent against a severe second peak.

### Maximally severe restrictions indicate counter-intuitive consequences: COVID-19 “eradication” red herring

Our model indicates that maximally severe restrictions (e.g., complete quarantine and isolation) would lead to unintended consequences and would, under most scenarios, lead to a severe second peak. Initially, maximally severe restrictions would appear to be effective in controlling the spread of COVID-19. However, these maximally severe restrictions would also leave the vast majority of the population in the “susceptible” group. Because of the characteristics and infectiousness of COVID-19, a severe second peak is likely to result if these restrictions are lifted at any point short of global eradication (unless vaccine becomes available or other mitigation methods are considered). Our model indicates that if maximally severe policies are lifted after 60 days, even though infection curves appear to suggest that the disease has been controlled and contained, a severe outbreak will likely occur, and this peak will likely far exceed health care capacity (see Figure 2A). A similar result occurs if the timeline is expanded to 90 days (see Figure 2B). Our model suggests that, in the absence of alternative mitigation measures, maximally severe restrictions is not an optimal strategy against COVID-19, even if it initially appears to be successful in reducing spread.

## Conclusion

Our model highlights the importance of a measured response to COVID-19 control. Social distancing policies are expensive and disruptive, and there is a desire and an urgency to return to normalcy and pre-pandemic levels of travel and contact. As social distancing measures are shown to be effective at reducing disease spread, we expect those pressures to increase. However, our model shows the dangers of lifting or relaxing social distancing policies too soon. The novelty of COVID-19 means that the susceptible population is uniquely large, and the highly infectious nature of COVID-19 means that even a minimal residual presence of the virus can lead to a second severe outbreak if social distancing policies are lifted or relaxed too quickly. Our model suggests that a longer-term initial period of restrictive social distancing, coupled with a gradual reduction in the severity of social distancing, would be more effective in controlling the COVID-19 outbreak.

## Data Availability

There is no data involved in this paper. However, there is an interactive notebook illustrating the model results in a dynamic way.

https://www.wolframcloud.com/obj/22f74499-3d57-49ed-b872-9fbf21f719eb

## Acknowledgments

ZF’s research is partially supported by NSF grant DMS-1814545.

## Disclaimer

The findings and conclusions in this report are those of the authors and do not necessarily represent the official views of the National Science Foundation.

